# A non-parametric mathematical model to investigate the dynamic of a pandemic

**DOI:** 10.1101/2020.04.30.20086199

**Authors:** Robert Sosa, Wilfredo Sosa

## Abstract

Populations are diverse in size, capacity response, and measures to contain a pandemic such covid19. Then, it rises serious and critical questions to whether general measures can be taken worldwide. Also, it is unclear if conventional parametric methods are suitable to study pandemics since their dynamic is modulated by biological, economical, environmental, social, and cultural factors. In this manuscript, we apply a recently developed non-parametric mathematical method that comes from regional economy, to investigate the dynamic of a pandemic. We apply this novel methodology to study the ongoing covid19 pandemic in all USA states and in the country itself. The generality of our methodology makes it suitable to investigate also the worldwide dynamics of diseases such HIV or tuberculosis.

## 1 Introduction

The ongoing pandemic caused by the severe acute respiratory syndrome coronavirus 2 (SARS-Cov2) has infected more than two million people and claimed more than 130,000 lives worldwide until 17 April 2020 [1]. Since neither effective antiviral drugs or vaccine exist in a new outbreak, preventive measures are needed to stop the spreading of virus. Therefore, control measures (e.g. social distancing) should be properly undertaken in order to modulate the trajectory of the epidemic and minimize the impact on the health care system [2]. However, it is not clear wether similar measures should be taken by regions with a different health care capacity response, population size, and the number of new positive along with fatal cases daily. It is also unclear which measures are more effective than others in different regions. These concerns are even more critical and urgent since new outbreaks are predicted in a not-to-far future [3].

Several factors may influence the trajectory of the pandemic which may difficult its modeling and thus model predictions become unreliable. First, for the case of coronavirus, two strains are identified: the S- and L-type, being the later the most aggressive and infectious. The L-type was more prevalent in early stages of the outbreak in Wuhan than the S-type, and the relative prevalence of these strains have changed during the pandemic [4]. We remark that it is unclear if the relative frequency between the S- and L-strain follows a similar pattern in different regions affected by the virus. Second, environmental factors such as humidity, temperature, and airflow are relevant to determine the transmission and the stability of human respiratory viruses such as the SARS-Cov2 [5], [6]. Therefore, the diversity of environmental conditions will also determine the dynamic of the pandemic in different regions affected by the virus. Third, risk factors such as older age or cardiac injury were associated with higher in-hospital mortality [10], [11]. This result indicates that the dynamic of the covid19 pandemic will be different in regions whose the elderly represents a higher percentage of the population. Fourth, the coronavirus spreads mainly through close interaction with infected people, by coming into contact with droplets, exhaled or coughed, that contain the coronavirus [18]. Thus, public measures are designed to reduce people’s mobility and so the spreading of the disease [2]. However, we remark that the severeness of these measures are associated to the impact on the economy and are culturally and socially influenced whereas its effectiveness depends on human behavior. Lastly, social inequality, poverty, malnutrition, the lack of clean water and adequate sanitation, and the lack of infrastructural and financial preparedness significantly increase individual susceptibility to infection, and increase the risk of morbidity-mortality during an outbreak [12]. Therefore, parametric models are unrealistic because they ignore or are not able to account for all these collective factors that may alter the dynamic of a pandemic [7], [8], [9]. In this line, non-parametric models will be more informative where parametric models fail to account for all these factors.

Here, we apply the recent modification of a well-established and a non-parametric methodology in regional economy, the location quotient (*LQ*) to understand the dynamic of a pandemic. The location quotient (*LQ*) was introduced by Robert Murray Haig in 1920 [13] and have a wide application in economy, [14], [16], [15] and [17]. However, *LQ* was seriously criticized. In regional economy, the *LQ* needs two dimensions for its definition, the economical dimension (productive sector of the economy) and the spacial dimension (the regions of the economy). Recently, in a PhD thesis^1^ at the Catholic University of Brasilia, the *LQ* was modified by considering an extra dimension, the time. With this modification, now the applicability of this non-parametric framework can be extended to problems dealing with a temporal dynamic, which is the case of the ongoing covid19 pandemic.

## 2 A new non-parametric model

Here *V*(*s,r,t*) is the observation of the variable *s* in the region *r* at time *t*. The new dimension *t*, define an horizon *H* for observations (*H* units of times). In this horizon, we define periods, where each period has *P* units of time, which define a total of *H* – *P* +1 periods. Each period *p* can be seen as a moving window of size *P* spanning the complete horizon *H*. In term of lineal Algebra, we fix a region *r* ∊ {1, 2, ···, *R*} and a period *p* ∊ {1, 2, ···, *H* – *P* + 1} to define a matrix with *S* rows and *P* columns, called the Regional Matrix of location quotients, denoted by 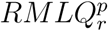. Here, each matrix has associated a semi definite symmetric matrix (the covariance matrix), denoted here as 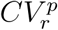. Since 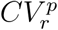 is symmetric and semi definite, it has real eigenvalues and eigenvectors. The interpretation of each eigenvalue λ is as follows: if *m* is the mean of the rows of the matrix 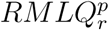 and *£* is the line that passes through *m* with direction of the eigenvector *v* associated to λ, then λ represent the dispersion of the projections of the rows of matrix 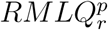 over *£*. So, all the eigenvalues represent the dispersion of the rows respect to *m*. Since the focus is the concentration of all the productive sectors in the region *r* at the period *p*, then the norm of the vector contains all the eigenvalues of the matrix 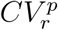 is defined as the concentration index of all productive sectors in the region *r* at the period *p*.

However, the *CI* matrix is not sufficient to understand the global behavior of each variable in the country, because each *CI* component in the matrix represents dispersion of the behavior of three variables in a given state at certain period. Therefore, the analysis of *CI* is complemented with the study of the global percentage *GP* to describe the global behavior of each variable *s* in the country at each time *t*. Therefore, considering both GP and *CI* we can investigate a global and local dynamic of the covid19 pandemic in different regions of the country and in the country itself.

## 3 Application of the non-parametric model to the covid19 pandemic

The pandemic of the covid19 is controlled by performing nation wide tests, the results to this test are positive or negative (the asymptomatic cases and the ones without the virus), and a portion of infected people (positive test) result in deaths. So, we consider the variables, the number of test *s* = 1, positive test *s* = 2 and deaths *s* = 3 as a productive sector of the covid19, for each state *r* in USA. The economy is USA (without regions with low number of data). Therefore, we have,

1. *R* = 52 (USA states or regions, it includes also Hawaii, Alaska, and Puerto Rico)
2. *S* = 3 (the three variables)
3. *H* = 41 (from March 06 to the date the data was collected, April 16)
4. *P* = 7 time spam of the government response to take measures (“moving window”). For a first approximation, we chose seven days.

So,

1. *V*(*s, r, t*) component of tensor *V* with size *S* × *R* × *H* (the data),
2. 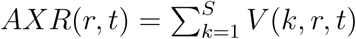 component of aggregation by regions matrix *AXR* with size *R* × *H*,
3. 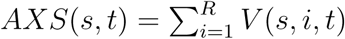 component of aggregation by sector matrix *AXS* with size *S* × *H*,
4. 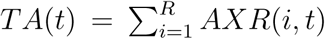 component of total aggregation vector *TA* with *H* components,
5. *GP*(*s, t*) = *AXS*(*s, t*)*/TA*(*t*) component of global percentage matrix *GP* with size *S* × *H*,
6. *LP*(*s, r, t*) = *V*(*s, r, t*)*/AXR*(*r, t*) component of local percentage tensor *LP* with size *S* × *R* × *H*.

So, the location quotient of the variable *s* in the state *r* at time *t* is defined as:

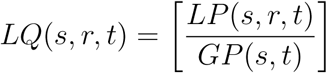

The matrix *CI* is built from the *LQ* tensor for each state at each period. The tensor *LQ* is the transformation of the data *V*. So, *CI* has *R* rows and *H* – *P* +1 columns. Each component *CI*(*r,p*) is defined as the norm of the vector that contains all the eigenvalues of the covariance matrix of 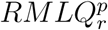 (here, 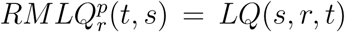 for *s* ∊ {1, 2, 3} and *t* ∊ {*p, p* +1, ··· *,p* + *P* – 1}). When *CI* (*r, p*) is close to zero, it means that each component *LQ*(*s, r, t*) of 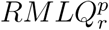 is close to one. In other words, the sequences of *LP*(*s, r, t*) values follows the dynamic of the corresponding part of the sequences of *GP*(*s, t*) values. However, when *CI*(*r, t*) is far from zero, then *LQ*(*s, r, t*) is far from one. It implies that *LP*(*s, r, t*) > *GP*(*s, t*) or *LP*(*s, r, t*) < *GP*(*s, t*).

## 4 The global view of the covid19 pandemic

In this section we start with the analysis of global percentage *GP*. Here, the pandemic is controlled when there is no new death ({*GP*(3, *t*)} converges to zero) and there is not new positive test in the country ({*GP*(2, *t*)} converges to zero). This indicates that {*GP*(1, *t*)} converges to one, since *GP*(1, *t*) + *GP* (2, *t*) + *GP* (3, *t*) = 1.

Daily historical data of number of tests, positives tests, and deaths were collected from the database “COVID Tracking Project^2^” for all USA states. After collecting these data, we applied our model using a code written in Scilab and built the matrices *GP* and *AXS* for USA. We mentioned earlier that the coronavirus mainly spreads through close interaction with infected people [18]. As a result, the Centers for Disease Control (CDC) and all states of USA have taken several measures (between march 15 - 16 +/ – 5 days, Figure 1) such legally enforcing social distancing, declaring state of emergency, or closing all non-essential businesses to limit people’s contact [18], namely reduce people’s mobility. Note that some measures are more flexible than others and they also differ between USA states, making it hard to correlate them with *GP* and *AXS*. Therefore, we also collected people’s mobility daily data of all USA states to correlate them with the the corresponding *GP* and *AXS* dynamics (Figure 1). We believe that the reduction of people’s mobility is the result of all types of diverse measures taken by a particular USA state or the country to reduce the spread of the coronavirus. In other words, restriction of people’s mobility aims to the convergence *GP*(2, *t*) and *GP*(3, *t*) to zero. Daily historical people’s mobility data per USA state was collected from the ”wallethub” database (https://wallethub.com/edu/).

**Figure 1:**
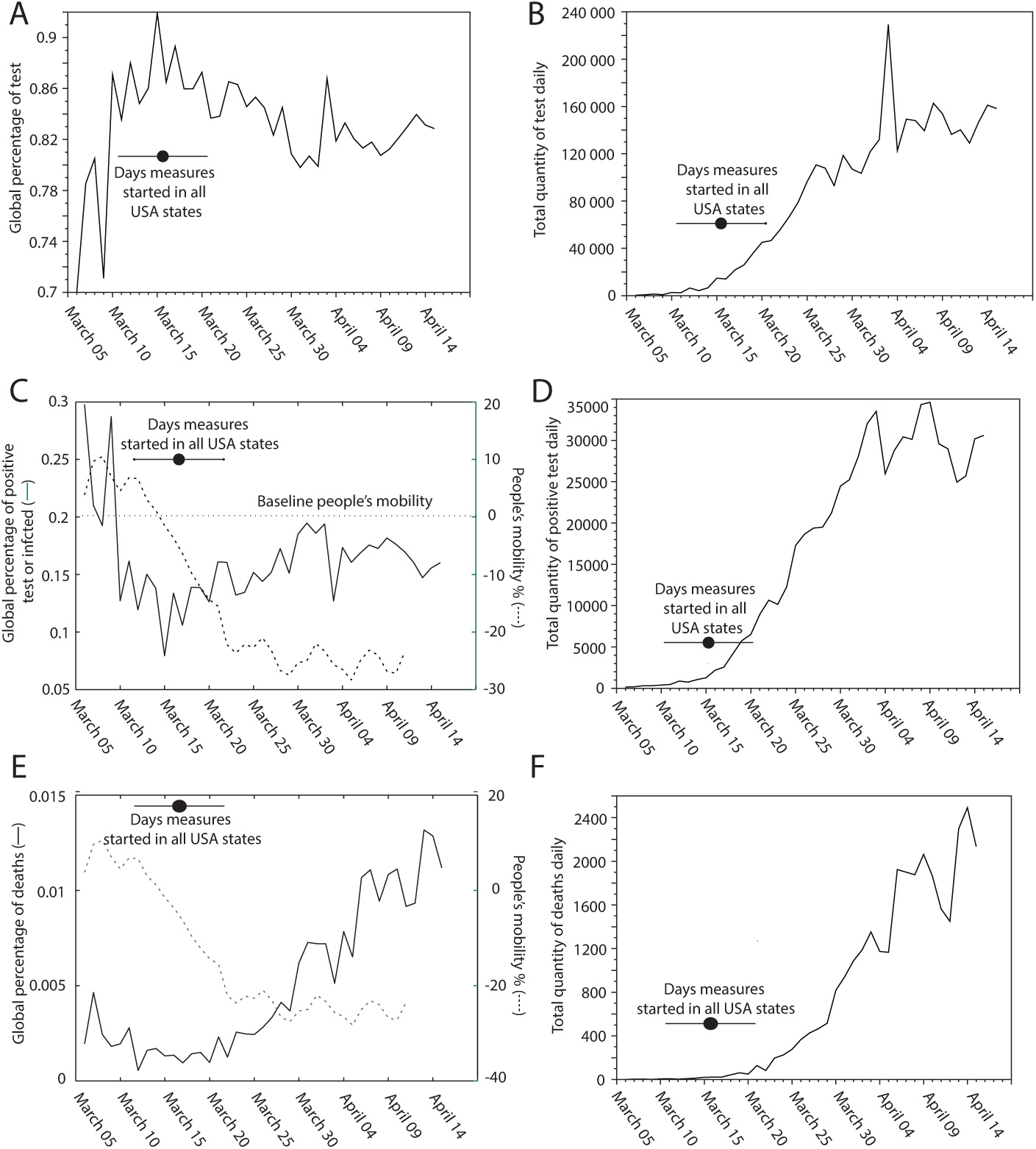
The global behavior of the covid19 pandemic: (A) Daily behavior of Global percentages of tests {*GP*(1, *t*)}. (B) Total quantity of tests *AXS*(1, *t*) daily. (C) Daily behavior of Global percentages of positives test {*GP*(2, *t*)}. (D) Total quantity of positives test *AXS*(2*, t*) daily. (E) Daily behavior of Global percentages of deaths {*GP*(3*, t*)}. (F) Total quantity of daily deaths *AXS*(3,t). Note that here at any given time t, {GP(1, t)} + {*GP*(2, *t*)} + {*GP*(3*, t*)} = 1. Continuous black lines refer to the experimental temporal dynamic of {*GP*(1, *t*)}, {*GP*(2, *t*)}, and {*GP*(3, *t*)} values from March 05 to April 16. Discontinuous black line refers to people’s mobility from March 05 to April 11. The horizontal black line depicts the average date (dot) and the standard deviation (bar) of the distribution of USA states when they start public measures, 15 - 16 +/ – 5 days.

From our results (Figure 1), the following observations are pointed out:

1. Within the first ten days of the pandemic, from March 06 to march 16 of the pandemic, the global percentage of positive test ({*GP*(2, *t*)}) approached to zero (from 0.3 to 0.1) whereas the global percentage of deaths ({*GP*(3, *t*)}) remained close to zero (around 0.002) (Figure 1C, 1E). Consequently, the global percentage of test ({*GP*(1)}) approached to one or remained close to one (Figure 1A). These observations indicate that the positive test (infected people) and deaths were close to be contained ({*GP*(2, *t*)}, Figure 1C). Note that the reduction of {*GP*(3, *t*)} and {*GP*(2, *t*)} is accompanied by the decrease of people’s mobility from around 10 to 0%, where public measures start taking place between march 15-16 (Figure 1E). Note that the aggregation of positive test ({*AXS*(2, *t*)}) and fatal cases ({*AXS*(3, *t*)}), increased monotonically during 29 day (from March 06) (Figure 1D, 1F).
2. After March 16, the global percentage of positive test ({*GP*(2, *t*)}) slightly increased (until march 26) but then remained stationary, oscillating around 0.16 for the rest of the pandemic. Meanwhile, the global percentage of fatal case ({*GP*(3, *t*)}) still remained close to 0.002 before March 26. Coincidentally, when people’s mobility decreases to around -25%, the ({*GP*(2, *t*)}) remained stationary. This observation suggests that the spread of the virus is contained when a public measures result in a significant decrease of more than 25% of people’s mobility. There is a time interval of around 13 days where people mobility goes from +5 to -25% (Figure 1). Therefore, if public measures were taken at least 13 days before (between march 2 - 3), the covid19 pandemic could have been controlled ({*GP*(2, *t*)}) close and approaching to zero).
3. Since March 31, the global percentage of fatal cases ({*GP*(3, *t*)}), increased monotonically from 0.002 to below 0.014. This result indicates that everyday more infected people is dying, namely the fatality rate is worsening in USA (Figure 1E).

It is possible that before March 16, the covid19 pandemic was close to be contained since the hospital capacity response (e.g. ICU beads per 10000 people) was not overwhelmed. However, after March 26, the hospital capacity response may be surpassed by the increasingly number of infected people (Figure 1D). In USA, around 68,000 people die annually due to the lack of health care insurance [19]. Thus, uninsuranced people may also contribute to the increase of the fatality rate, as a consequence of the lack of treatment. We remark that the behavior of {*GP*(2, *t*)} and {*GP*(3, *t*)} within the first ten days, where the pandemic was close to be contained, cannot be easily observed classical methods such counting (or accumulating) the new the number of infected people (positive tests) and deaths daily (Figure 1D, 1F).

The {*GP*(2, *t*)} and {*GP*(3, *t*)} values define the global dynamic of the covid19 pandemic in the country, USA is this case. If they move away from zero, then it is necessary to establish more restrictive measures to people’s mobility to reduce the spreading of the virus. Therefore, federal public measures should be established and their effectiveness should be monitored based on the periodic analysis and the behavior of {*GP*(2, *t*)} and {*GP*(3, *t*)}. Now, we cannot assume all states are equally successful in controlling the spreading and the lethality of the pandemic, in fact some USA states may be contributing negatively or positively to the global dynamic. To detect which USA state has a negative contribution to *GP*, we then proceed with the analysis of the concentration indexes *CI* and the local percentage *LP* (next section).

## 5 The local view of the covid19 pandemic

In this section we will be focused on the local dynamic in USA states and their contribution to the global behavior of the pandemic in the country, considering the matrix *CI* and the tensor of local percentage *LP*. The matrix *CI* describe the behavior of the dispersion of these *LP* values, relative to *GP*, for a particular state *r* within a certain period *p*. For each state *r*, the sequence {*CI*(*r*, *p*)} is close to zero if the state *r* follows the global trend of the global percentage at one given period *p*, namely {*LP*(*s*, *r, t*)} and {*GP*(*r, t*)} are very similar. On the other hand, a large {*CI*(*r*, *p*)} value (away from zero) indicates that this particular USA state *r* will contribute negatively or positively to the global percentage at one given period *p*. For the case of deaths and infected people, a negative contribution is when *GP*(*s, t*) *< LP*(*s, r, t*) and a positive contribution is when *GP*(*s, t*) > *LP*(*s, r, t*).

Through the individual analysis of *CI* and *LP* values of all USA states, we carefully selected the ones that have high *CI* values (more than 1) and *GP*(*s, t*) < *LP*(*s, r t*) for the case of positive test (infected people, *s* = 2) and deaths (*s* = 3). After this process, we selected six USA states for the case of positive test: Alabama, California, Maine, Mississippi, New Jersey, New York, Oklahoma, and Washington (Figure 2C). For the case of deaths we selected: Maine, Michigan, New Jersey, and New York (Figure 2D). Some of USA states, have high *CI* values than the ones selected, either at the beginning, at the end or in a middle period, but *GP*(*s, t*) – *LP*(*s, r, t*) was fluctuating around zero, indicating that on average they follow the global trend. The other USA states have *CI*(*r, p*) near to zero along with nearly zero or positive *GP*(*s, t*) – *LP*(*s, r, t*) values through the duration of the pandemic. These USA states were also not selected for further analysis. To rationalize our findings with factors that may be critical for the increase of the concentration of infected people and deaths in each USA state. Here we consider: the economic accessibility to test and treatment (measured as percentage of people in poverty and without health care insurance), the hospital capacity response to treat infected people and prevent deaths (ICU beads per 10000 people), the propensity of people to get infected (measured as population density, and people’s mobility), and people in high risk (percentage of people of 65 years older and above).

**Figure 2:**
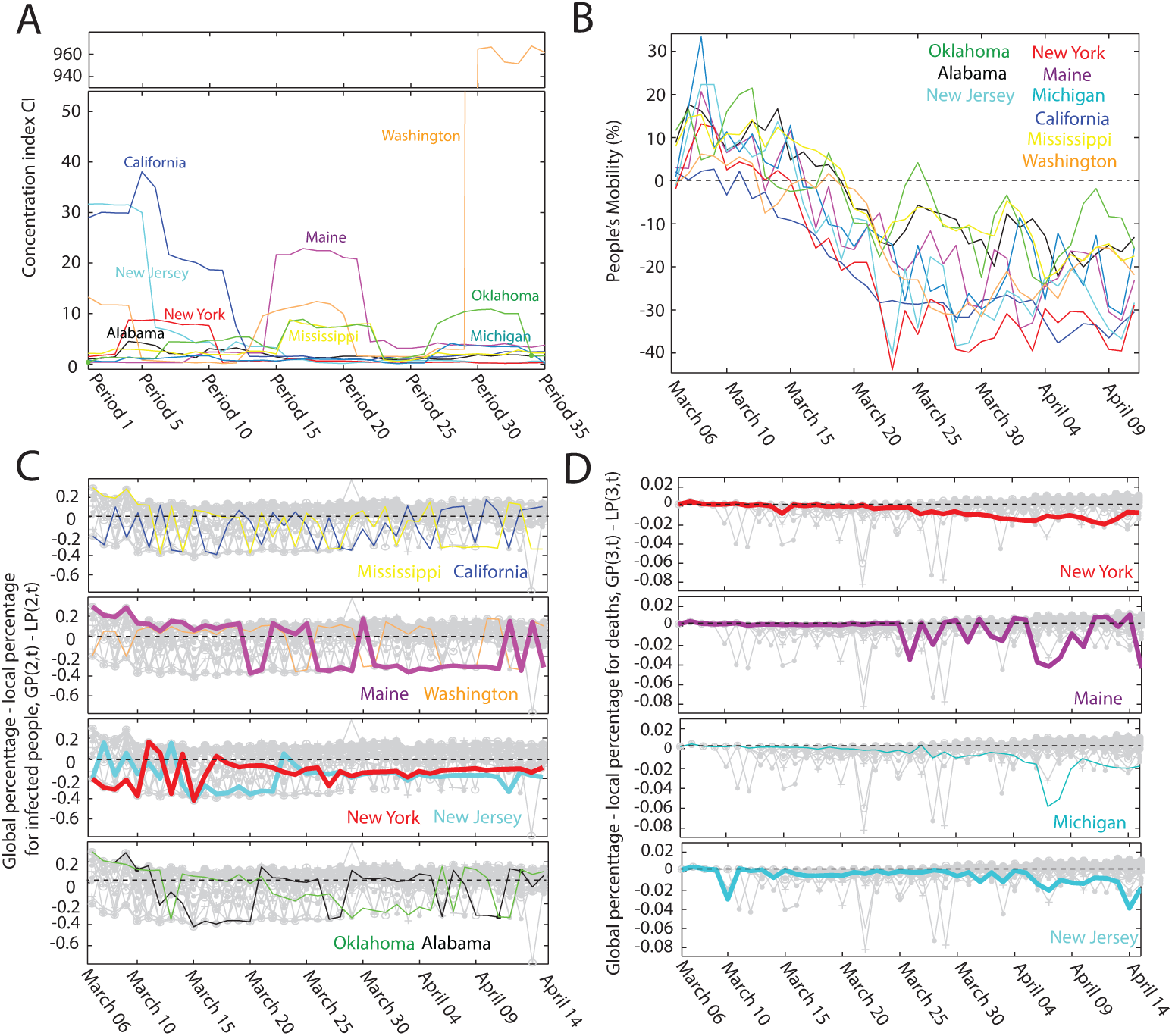
The local dynamic of the covid19 pandemic. (A) Dynamic behavior of CI ¿ 1 values for 7 states in the 35 periods: Alabama (black), California (dark blue), Maine (purple), Michigan (stone blue), Mississippi (yellow), New Jersey (sky blue), New York (red), Oklahoma (green), and Washington (orange). (B) Comparative analysis of people’s mobility profile. (C) *GP*(s, *t*) – *LP*(*s, r, t*) profile of positive test (infected people). (D) *GP*(*s, t*) – *LP*(*s, r, t*) profile of deaths. For the case of (B), (C) and (D), the dynamic is depicted from March 05 to April 11 for the seven USA states. The grey background, in (C) and (D), is the GP(s,t) – *LP*(*s, r, t*) profile of all USA states that are not considered in the analysis in (A), since they follow the global trend *GP*(*s, t*) = *LP*(*s, r, t*) or contribute positively to the global trend *GP*(*s, t*) > *LP*(*s, r, t*)

From our results (Figure 4), the following observations are pointed out:

1. Only three states are the ones that contribute negatively to the global trend of positive test (infected people) and deaths. They are New Jersey, New York, and Maine (Figure 2C, 2D). For these USA states, the local dynamic of the covid19 pandemic is worsening in New Jersey and Maine whereas New York seems to approach to the global trend (Figure 2C, 2D). Then, our model suggest that a more drastic reduction of people’s mobility are needed to further contain the pandemic in these states. Remarkably, this prediction is consistent with the fact both New York and New Jersey together account for around 42% of the total infected people and deaths in USA according to the CDC data.
2. Note that the state of Michigan is following the global trend of infected people, indicating that public measures are taking effect (Figure 2C). However, the fatally rate is significantly higher than the global percentage and also seem to be worsening (Figure 2D). Consistently, Michigan is already ranked third in number of deaths in USA.
3. There are five USA states whose fatality rate follows or is below the global trend but the local percentage of infected people seem to be worsening. They are Alabama, California, Mississippi, Oklahoma, and Washington.

It is not clear wether the earlier the measures were taken, the population density, the lack of health care insurance, poverty, (Figure 3A-C) or the relative magnitude of reduction of people’s mobility (Figure 2B) contribute significantly to the relative reduction in the spreading of the virus in USA states. For example, California achieved earlier (since March 11) and more dramatic reduction of people’s mobility (down to -35%) whereas Michigan reduced people mobility around 4 days later and only down to -30% (Figure 2B). However, Michigan was more successful to contain the spreading of the virus. We mentioned earlier that the reduction of people’s mobility aims to minimize people’s contact to stop the spreading of the virus [18]. Therefore, if the local percentage of infected people increases or it is still higher compared to the global trend, then more severe public measures are needed to limit people’s contact even further.

**Figure 3:**
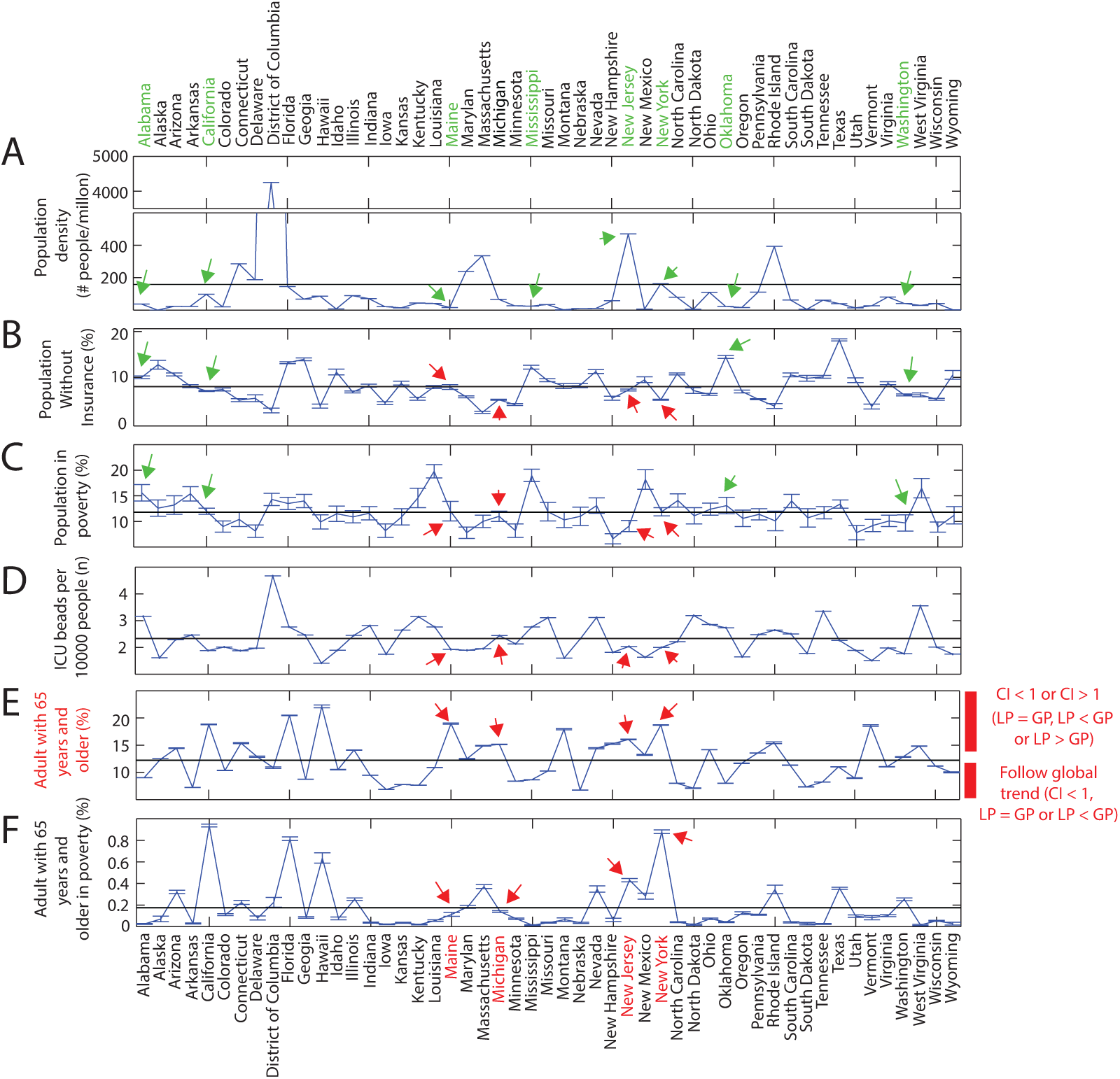
Factors that may contribute to the dynamic of the pandemic in all USA states: (A) Population density, (B) % of people without insurance, (C) % of people in poverty, (D) Number of ICU beads per 10000 people, (E) % of population with 65 years old and above, and (F) % of population with 65 years old and above in poverty. The states highlighted in red (bottom) refer to the ones in Figure 2D. The states highlighted in green (top) refer to the ones in Figure 2C. Green and red arrows help to the eyes to indicates the corresponding value of the USA states selected in the analysis. Average values are depicted by the horizontal line in each graph. Note that all USA states with high mortality can be explained by their higher % of population with 65 years old and above (highlighted in red) than the average. Above the average, USA states have either *CI >* 1 or *CI <* 1, whereas above the line, they have necessarily *CI <* 1 (Figure 3E, right)

Risk factors such as older age is associated with higher in-hospital mortality [10], [11]. Also, the lack of infrastructure such ICU beads contributes to the increase of the fatality rate [12]. Then, the high mortality in New York and New Jersey can be explained as follows i) both have 53% more than the average of a the percentage of adult population above 65 years, and ii) 115 and 340% more of the percentage of adult population above 65 years in poverty than the average in USA, respectively (Figure 3E-F). Only for the case of New York, this high fatality rate can also be associated with the 13% less ICU bead per 1000 people than the average (Figure 3C). For the case of Michigan and Maine, the high mortality can be explained only by the higher percentage of adult population above 65 years than the average in USA, around 24 and 56% respectively (Figure 3E-F). Only for the case of Maine, the high mortality can also be associated with the 16% less ICU bead per 1000 people than the average (Figure 3C). Overall, we conclude that a USA state with high mortality can be associated to a higher percentage of adult population above 65 years relative to the average. Other factors such the lack of ICU beads and the poverty of this high risk population can also explain the high mortality relative to the global trend, but they are not Determinant. Nonetheless, these factors do not necessarily imply that a particular USA state will have a high fatality rate. For instance, Florida has 66 and 307% more than the average of the global percentage of adult population above 65 years and in poverty, respectively; and yet the mortality rate in this case follows the trend of the global percentage. On the other hand, a lower percentage of these high risk population does necessary implies a lower fatality rate (Figure 3E), which is the case for all USA states that were not selected for the analysis.

To the best of our knowledge, epidemiologist lack of a non-parametric model to study the dynamic of diseases or pandemics. In a broader view, parametric models ”force the data” to fit into a particular model with numerous assumptions that are unrealistic, since processes such a diseases (e.g. HIV or tuberculosis) or pandemics are influenced by biological, economical, environmental, social, and cultural factors. In this line, our non-parametric model offer a new perspective since it deals directly with the experimental data, and thus potentially provide more information that can be used as a background to take preventive and critical measures to reduce the world-wide negative impact of diseases such HIV, tuberculosis or the ongoing covid19 pandemic.

## Data Availability

Data can be provided upon request to the authors

## 6 Figures

## Footnotes

1 This thesis was advised by the second author, the PhD student is Jose França and the title is “Quocientes de lacação e politicas públicas de desenvolvimento regional: Teoria y aplicações”. the defense of this thesis was rescheduled due to the quarantine in Brasilia.

1 https://covidtracking.com/data

